# OpenSAFELY NHS Service Restoration Observatory 1: describing trends and variation in primary care clinical activity for 23.3 million patients in England during the first wave of COVID-19

**DOI:** 10.1101/2021.01.06.21249352

**Authors:** The OpenSAFELY Collaborative, Helen J Curtis, Brian MacKenna, Richard Croker, Peter Inglesby, Alex J Walker, Jessica Morley, Amir Mehrkar, Caroline E Morton, Seb Bacon, George Hickman, Chris Bates, Jessica Morley, David Evans, Tom Ward, Jonathan Cockburn, Simon Davy, Krishnan Bhaskaran, Anna Schultze, Christopher T Rentsch, Elizabeth Williamson, William Hulme, Helen I McDonald, Laurie Tomlinson, Rohini Mathur, Henry Drysdale, Rosalind M Eggo, Kevin Wing, Angel YS Wong, Harriet Forbes, John Parry, Frank Hester, Sam Harper, Stephen JW Evans, Ian J Douglas, Liam Smeeth, Ben Goldacre

## Abstract

**Background:** The COVID-19 pandemic has disrupted healthcare activity globally. The NHS in England stopped most non-urgent work by March 2020, but later recommended that services should be restored to near-normal levels before winter where possible. The authors are developing the *OpenSAFELY NHS Service Restoration Observatory*, using data to describe changes in service activity during COVID-19, and reviewing signals for action with commissioners, researchers and clinicians. Here we report phase one: generating, managing, and describing the data.

**Objective:** To describe the volume and variation of coded clinical activity in English primary care across 23.8 million patients’ records, taking respiratory disease and laboratory procedures as key examples.

**Methods:** Working on behalf of NHS England we developed an open source software framework for data management and analysis to describe trends and variation in clinical activity across primary care EHR data on 23.8 million patients; and conducted a population cohort-based study to describe activity using CTV3 coding hierarchy and keyword searches from January 2019-September 2020.

**Results:** Much activity recorded in general practice declined to some extent during the pandemic, but largely recovered by September 2020, with some exceptions. There was a large drop in coded activity for commonly used laboratory tests, with broad recovery to pre-pandemic levels by September. One exception was blood coagulation tests such as International Normalised Ratio (INR), with a smaller reduction (median tests per 1000 patients in 2020: February 8.0; April 6.2; September 7.0). The overall pattern of recording for respiratory symptoms was less affected, following an expected seasonal pattern and classified as “no change” from the previous year. Respiratory tract infections exhibited a sustained drop compared with pre-pandemic levels, not returning to pre-pandemic levels by September 2020. Various COVID-19 codes increased through the period. We observed a small decline associated with high level codes for long-term respiratory conditions such as chronic obstructive pulmonary disease (COPD) and asthma. Asthma annual reviews experienced a small drop but since recovered, while COPD annual reviews remain below baseline.

**Conclusions:** We successfully delivered an open source software framework to describe trends and variation in clinical activity across an unprecedented scale of primary care data. The COVD-19 pandemic led to a substantial change in healthcare activity. Most laboratory tests showed substantial reduction, largely recovering to near-normal levels by September 2020, with some important tests less affected. Records of respiratory infections decreased with the exception of codes related to COVID-19, whilst activity of other respiratory disease codes was mixed. We are expanding the *NHS Service Restoration Observatory* in collaboration with clinicians, commissioners and researchers and welcome feedback.

## Background

The ongoing pandemic due to severe acute respiratory syndrome coronavirus 2 (SARS-CoV-2) has affected over 70 million people worldwide with at least 1.6million deaths due to COVID-19 as of December 2020[1]. The need to direct resources towards patients requiring treatment for COVID-19, and to minimise opportunities for spread by reducing face-to-face contact between individuals, meant that routine healthcare services faced significant levels of disruption. Shortly after declaring COVID-19 a *Public Health Emergency of International Concern*, the World Health Organisation (WHO) reiterated their operational preparedness guidance. The intention of the guidance was to provide countries with advice on how to minimise direct and indirect mortality from COVID-19 through the continued provision of essential services. Recommendations included rapid assessment of healthcare capacity, and the development of key performance metrics, and also highlighted the importance of keeping this data up to date [2,3]. The NHS in England responded to the emerging pandemic by stopping non-urgent work in hospitals, and suggesting that, where possible, patients should have non-urgent primary care appointments remotely [4].

In a rapid assessment conducted in May 2020, the WHO found that across the world the treatment of people with non-communicable disease (NCDs, non-infectious diseases not passed from person to person) had been considerably impacted by the severe disruption to the delivery of national healthcare services [5]. Subsequently, NHS England issued guidance on the “third phase” of the NHS response to COVID-19 on July 21st. One of the many recommendations was to restore NHS services to near-normal levels where clinically appropriate before winter, whilst remaining vigilant for a second wave [6].

OpenSAFELY is a new secure analytics platform for electronic patient records built by our group on behalf of NHS England to deliver urgent academic and operational research during the pandemic [7]: analyses can currently run across all patients’ full raw pseudonymised primary care records at 40% of English general practices, with patient-level linkage to various sources of secondary care data; all code and analysis is shared openly for inspection and re-use. A stated aim of OpenSAFELY is to assess “COVID Aftershocks” where we monitor data to measure and mitigate the indirect health impacts of COVID-19[8]. In order to produce the best possible insights across a range of diverse topics using this huge volume of activity and data, we are working with NHS England to create a programme of work that we have titled the *OpenSAFELY NHS Service Restoration Observatory*. Traditionally researchers using electronic health records data create bespoke manually curated “codelists” to identify certain diseases or units of healthcare activity. However as the scale of raw data is unprecedented, we are initially deploying a data-driven approach utilising natural hierarchies contained within CTV3 code structures (box 1). The insights generated will then be manually reviewed and prioritised by groups of clinicians and commissioners for further analysis. We aim to rapidly identify all important changes in clinical practice that have been collaboratively determined by clinicians and commissioners to be of high clinical importance, and then prioritise each relevant activity change for either remedial activity, additional monitoring, feedback to practices and regions, or further exploration.

In this paper we set out the first phase of this work, describing trends and variation in clinical activity codes to evaluate NHS service restoration from the first wave of the pandemic in England. We selected two clinical topic areas: “respiratory disease”, because this encompasses infectious diseases and common NCDs such as asthma and chronic obstructive pulmonary disease (COPD); and “laboratory procedures” including blood tests, because these are required in diagnosis and monitoring for a broad range of NCDs. We also developed a classification system to filter time trend data to identify changes of interest.

## Methods

### Study design

General practice clinical activity was described by conducting a retrospective cohort study using raw data from English NHS general practices.

### Data Source

Primary care records managed by TPP were analysed through OpenSAFELY, a data analytics platform created by our team on behalf of NHS England to address urgent COVID-19 research questions (https://opensafely.org). OpenSAFELY provides a secure software interface allowing researchers to run statistical analysis code across pseudonymised primary care patient records from England in near real-time within the electronic health record (EHR) vendor’s highly secure data centre, avoiding the need for large volumes of potentially disclosive pseudonymised patient data to be transferred off-site. This, in addition to other technical and organisational controls, minimises any risk of re-identification. Pseudonymised datasets from other data providers are securely provided to the EHR vendor and linked to the primary care data. The dataset contains information on 23.8 million people registered with GP surgeries using TPP SystmOne software as at 30 September 2020. It includes pseudonymised data such as coded diagnoses, medications and physiological parameters; no free text data are included. Practices are identified by pseudonymised codes only. Further details can be found under <u>information governance and ethics</u>.

### Study population

We included all patients registered with any practice using TPP EHR software (those with a registration beginning on or before 30 September 2020, which was still live until at least this date). All *coded events* between January 2019 and the end of September 2020 for this cohort were included. Coded events cover clinical diagnoses, symptoms, observations, investigations, administrative activities and other information recorded about patients. Codes may be manually entered by GPs/nurses or other practice staff, generated automatically when certain activities are carried out such as completing forms or templates, or derived from external sources such as secondary care.

### Data processing

Data was grouped at the practice level. We used each patient’s latest practice (as at 30 September 2020) as their assigned practice for all activity throughout the study period. We employed a data-driven approach in order to capture the most common coded events in primary care. We ranked codes according to the number of total occurrences in January-September 2020, excluding those with fewer than 1000. The total population in each practice was calculated as the total registered patients at 30 September 2020 and the same value used for every month.

### Clinical Code Classification

EHR systems in UK primary care have historically used a number of different clinical terminologies, including Read codes Version 2 (V2) and the Clinical Terms Version 3 (CTV3-box 1). Following the issue of a recent NHS Standard, all primary care systems must now be compliant with SNOMED CT [9]. More specifically, GP systems use a specific reference subset of the whole SNOMED CT terminology, known as the “GP Subset”. This subset very closely mirrors CTV3, the terminology historically used by TPP systems. There is a comprehensive, accurate mapping table between CTV3 and the UK “GP Subset” of SNOMED CT. Users of TPP GP EHR software default to work in SNOMED CT, but can choose to view records and browse codes using CTV3, in order to facilitate their transition to the new terminology. For the OpenSAFELY-TPP analysis here, we have worked within the CTV3 framework, in order to work rapidly and utilise its hierarchy structures.

CTV3 codes often give great detail on specific clinical findings or diagnoses. We therefore classified codes into a number of groups in order to ascertain more general trends. This was achieved in two ways. Firstly, CTV3 supports a comprehensive parent-child concept hierarchy, with defined relationships between different clinical concepts (box 1). We have utilised this hierarchy to support the classification of codes into high-level groups. Secondly, we supplemented these groups using the broad historical “category” structure of older terminologies, where parent-child relationships can be ascertained by a common set of initial characters. We acknowledge that this second approach can produce a small number of inappropriate or missing codes. It has, however, allowed us to work very rapidly to identify general trends. This is discussed further in key strengths and weaknesses

As an example, we grouped individual CTV3 codes into high-level topics using their first two digits, e.g. 42, “Haematology”. Secondly, we grouped codes by their first three digits to give a further breakdown of activities (e.g. 42Q, “Blood coagulation test”). Although this approach generally produces logical groupings, there may be some examples where this is not the case; for example “Asthma” (H33..) is grouped under “Chronic obstructive lung disease” (H3…), now considered an outdated classification. Further, these groups are non-exhaustive, because some codes related to these topics do not fall under this natural hierarchy; these ungrouped CTV3 codes (most commonly beginning with “X” or “Y”) are therefore presented individually.

#### Box 1

***Clinical Terms Version 3 - CTV3 codes***

CTV3 is a comprehensive computerised coding dictionary used by clinicians to record key clinical information about patients and also associated tests, diagnosis, medicines. CTV3 codes are used in the OpenSAFELY-TPP implementation, and fully align with the GP subset of SNOMED CT, the NHS standard [9]. There are almost 300,000 codes in total, each five characters long. CTV3 codes can be organised into hierarchies much like a book with chapters. CTV3 has a tree data structure with “parent” concepts describing broader clinical areas and “child” codes of increasing specificity as you move down the hierarchy. For example a concept such as “laboratory procedures” will have “child” codes that are more specific such as “haematology”, which will in turn be broken down further into increasingly detailed concepts. Most child codes can only have a single parent, although 3% have multiple parents.

### Code selection

Codes and groups were mapped to high level “concepts” (box 1) to assist with categorising them into related topics. We selected codes for each topic using concepts and/or keyword searches of code descriptions. For each topic we selected up to 75 of the most commonly occurring codes.

- *Pathology:* We identified codes under the following concepts: “Laboratory Procedures” (“4…”), “Laboratory test” (“X7A0B”) and “Laboratory test observations” (“X76sW”).
- *Respiratory:* We identified codes based on the keywords “respiratory”, “asthma”, “COPD”, “chronic obstructive”, “inhaler”. We did not specifically include keywords related to COVID-19, so it is likely that some but not all COVID-19 codes will be captured.

### Missing data

Missing data may arise through lack of inclusion of patients who have died; but this is likely to only have a limited impact on most codes, except in used for end-of-life issues which are relatively rare. We do not include patients who have moved away from TPP practices but we do include those who moved into their latest practice during the study period. Certain codes may appear to have missing data due to variable code use between practices, through e.g. selection of valid alternatives, use of different tools or templates, differing practice administrative processes, or use of on-screen tools for which codes do not get recorded. We could not account for these but expect that they will be generally consistent within each practice over time.

### Study Measures

We calculated the monthly incidence of each code (or group of codes) per 1000 registered patients at each practice. We calculated the median and deciles across all practices each month for each code, after excluding practices which never used that code in the study period. We present the data in time trend charts. All charts were collaboratively reviewed by clinicians and researchers; selected charts are shown in the Results section to illustrate key patterns, and all charts are shown in the associated OpenSAFELY Github repository[10] with selected charts in the Appendix.

In the case of all grouped (“parent”) codes, we present a table of the top (up to) five “child” codes to illustrate examples of individual codes captured, and we plot time trends of the top most common code. Fewer than five child codes being shown indicates that fewer than five codes exist in the group, or fewer than five reached the 1,000 minimum 2020 activity threshold.

### Classification of service restoration

With each chart we display the median value and interdecile range (IDR) for February, April and September 2020. April was identified as the first full month after full lockdown whilst September was the latest full month at the time of the initiating this analysis and before the “second wave” of infections would have been expected to influence health services once again. To aid interpretation, we provide an approximate classification based on changes to the median compared to the same month the previous year, which we have defined as our “baseline” (box 2).

#### Box 2

***Service change classification***

- For April and September:
  - *no change*: activity remained within 15% of the baseline level;
  - *increase*: an increase of >15% from baseline;
  - *small drop*: a reduction of between 15% and 60% from baseline;
  - *large drop*: a reduction of >60% from baseline.
- Overall classification:
  - *no change*: no change in both April *and* September;
  - *increase*: an increase in either April *or* September;
  - *sustained drop:* a small or large drop in April which has *not* returned to within 15% of baseline by September 2020;
  - *recovery*: a small or large drop in April, which returned to within 15% of baseline (“no change”) by September 2020.

### Software and Reproducibility

Data management was performed using Python and the OpenSAFELY software, with data extracted via SQL Server Management Studio and analysis carried out using Python. All of the code used for data management and analyses is openly shared online for review and re-use https://github.com/opensafely/restoration-observatory-intro-notebook.

### Patient and Public Involvement

This analysis relies on the use of large volumes of patient data. Ensuring patient, professional and public trust is therefore of critical importance. Maintaining trust requires being transparent about the way OpenSAFELY works, and ensuring patient voices are represented in the design of research, analysis of the findings, and considering the implications. For transparency purposes we have developed a public website which provides a detailed description of the platform in language suitable for a lay audience; we will be co-developing an explainer video; and we have presented at a number of online public engagement events to key communities. To ensure the patient voice is represented, we are working closely with appropriate medical research charities. In this instance, the draft paper has been shared with the Association of Medical Research Charities for general comment via a webinar and online feedback form, and specifically to Asthma UK and the British Lung Foundation for feedback from the perspective of those most affected by our findings, prior to publication in a peer reviewed journal.

## Results

Our study included 23,878,341 registered patients across 2,546 practices. Between 38.8%-100% practices were represented in each chart. From January-September 2020 for laboratory procedures we identified 83.6 million recorded events grouped under two-digit “parent” codes, and 235.7 million events grouped at three digits where possible, or at the full code level. By comparison, for respiratory disease we identified 1.4 million events at the two-digit parent code level, and 22.3 million events grouped at three digits where possible, or at the full code level. All charts are shown in the associated OpenSAFELY Github repository, with selected highlights below and in the Appendix.

### Pathology

Most two-digit high-level codes showed a similar pattern: a large drop (box 2) in activity in April 2020, recovering to normal levels over the summer. A representative example, haematology laboratory tests activity, is shown in Figure 1. The variation observed between practices in September (IDR 280.7) was similar to prior to the pandemic in February (IDR 276.8). More broadly, many individual tests had similar large drops in activity in April followed by substantial recovery, such as liver function, serum alkaline phosphate, serum creatinine and serum potassium (Appendix). The largest drop was observed in “Serum cholesterol (& level)” (figure 2) with the median falling by 90.2% from the previous year to 2.9 tests per 1000 in April. This recovered to near-normal levels by September, with the variation between practices similar to that seen in February (IDR 45.5 in both months). In contrast, blood coagulation tests such as INR, used to support management of anticoagulation, showed only a small drop in April (median 6.2/1000) compared to the pre-pandemic level (median 8.0/1000; figure 3).

**Figure 1:**
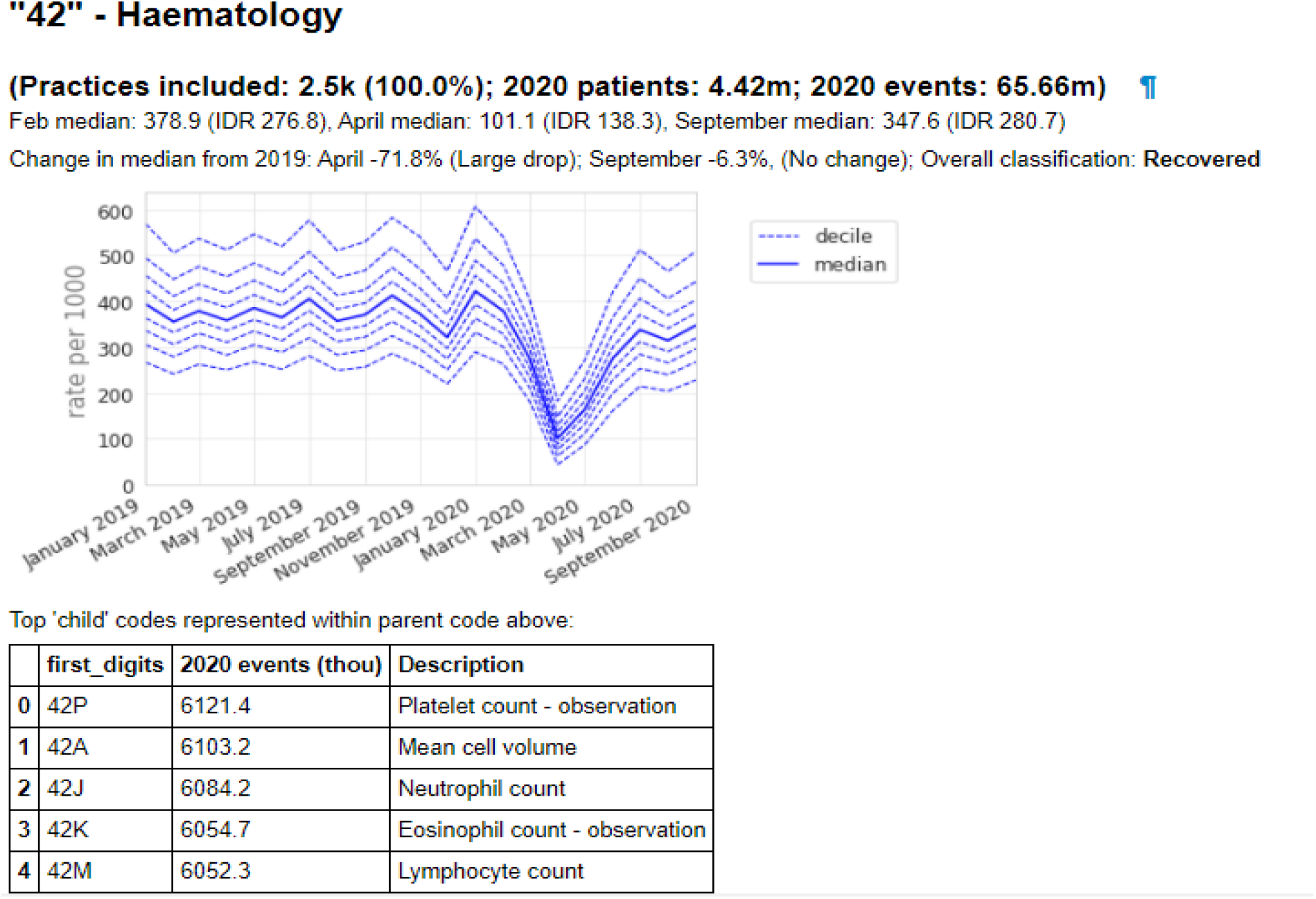
Recording of codes grouped under “Haematology” across TPP practices in England (January 2019-September 2020). The group includes CTV3 codes that begin with “42” and is not necessarily an exhaustive collection of every activity related to Haematology. The top 5 codes represented within this group are listed under the graph.

**Figure 2:**
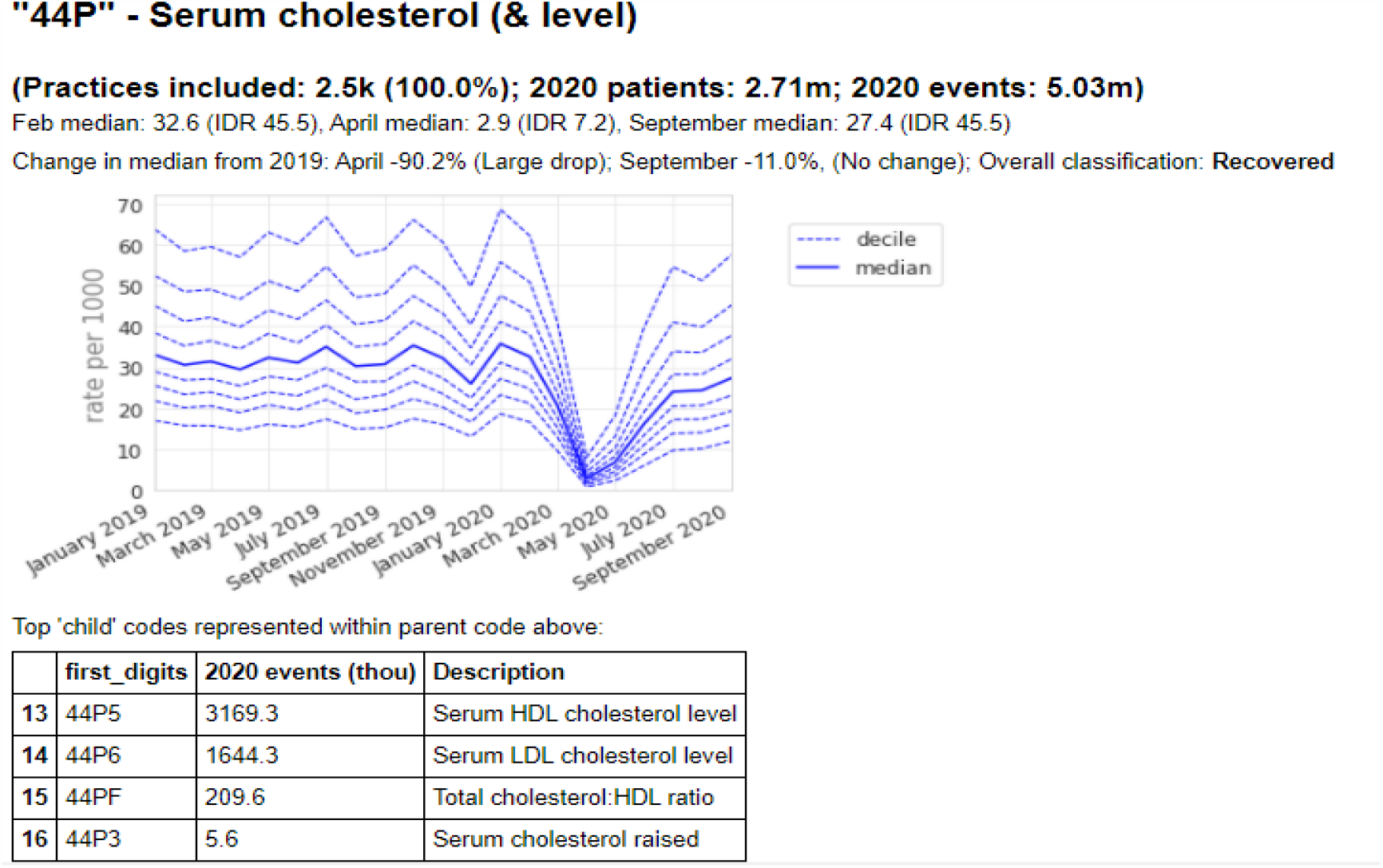
Recording of XE2eD (Serum cholesterol (& level)) across TPP practices in England (January 2019 - September 2020). This was the most common code we identified for this activity, but other codes may also be used to record the same or similar activity.

**Figure 3:**
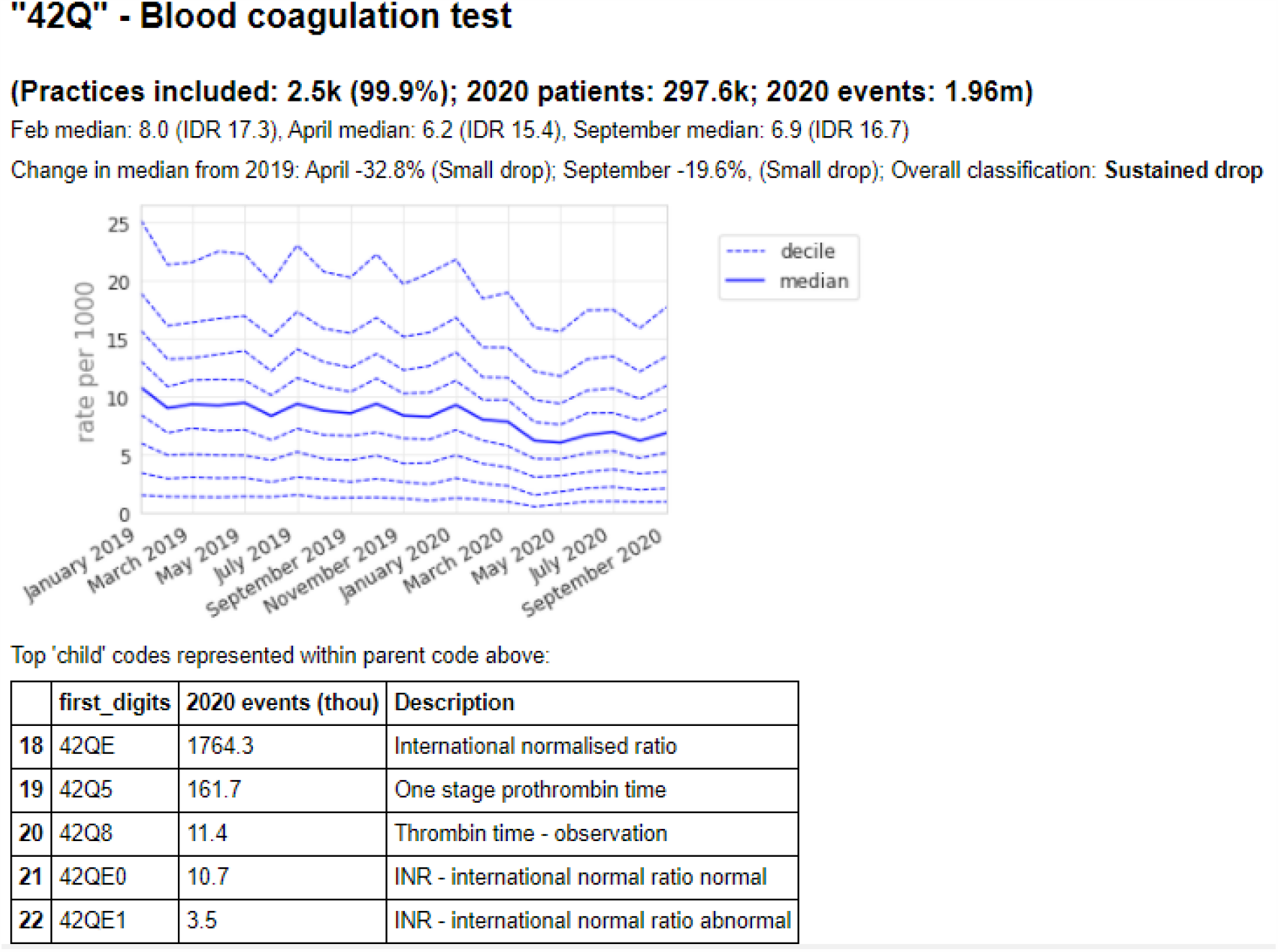
Recording of codes grouped under “Blood coagulation test” across TPP practices in England (January 2019 - September 2020). The group includes CTV3 codes that begin with “42Q” and is not necessarily an exhaustive collection of every activity related to Blood coagulation testing. The top 5 codes represented within this group are listed under the graph.

### Respiratory disease

The codes under respiratory disease can be broadly divided into those relating to symptoms and acute infections, and those relating to long-term conditions.

### Symptoms and acute infections

Recording of codes grouped under “respiratory symptoms” remained relatively stable throughout the observed period (figure 4), largely following an expected seasonal pattern. We selected three individual codes which demonstrate broad patterns in the recording of acute respiratory infections (figure 5). *Viral upper respiratory tract infection* (“Xa1sb”) exhibited a large and sustained drop compared with pre-pandemic levels, remaining substantially lower than the previous year (September −80.4%, figure 5a). Similarly, *Infection of lower respiratory tract* (“X1004”) experienced a large drop (−77.2%, figure 5b). As the recording of other respiratory infections began to drop, *Suspected coronavirus disease 19 caused by severe acute respiratory syndrome coronavirus 2* (“Y20cf”) increased rapidly during spring (April median 3.7/1000 IDR 6.8, figure 5c; other codes related to COVID-19 in the Appendix). This then decreased dramatically over the summer before increasing again in September (median 1.7/1000, IDR 2.8).

**Figure 4:**
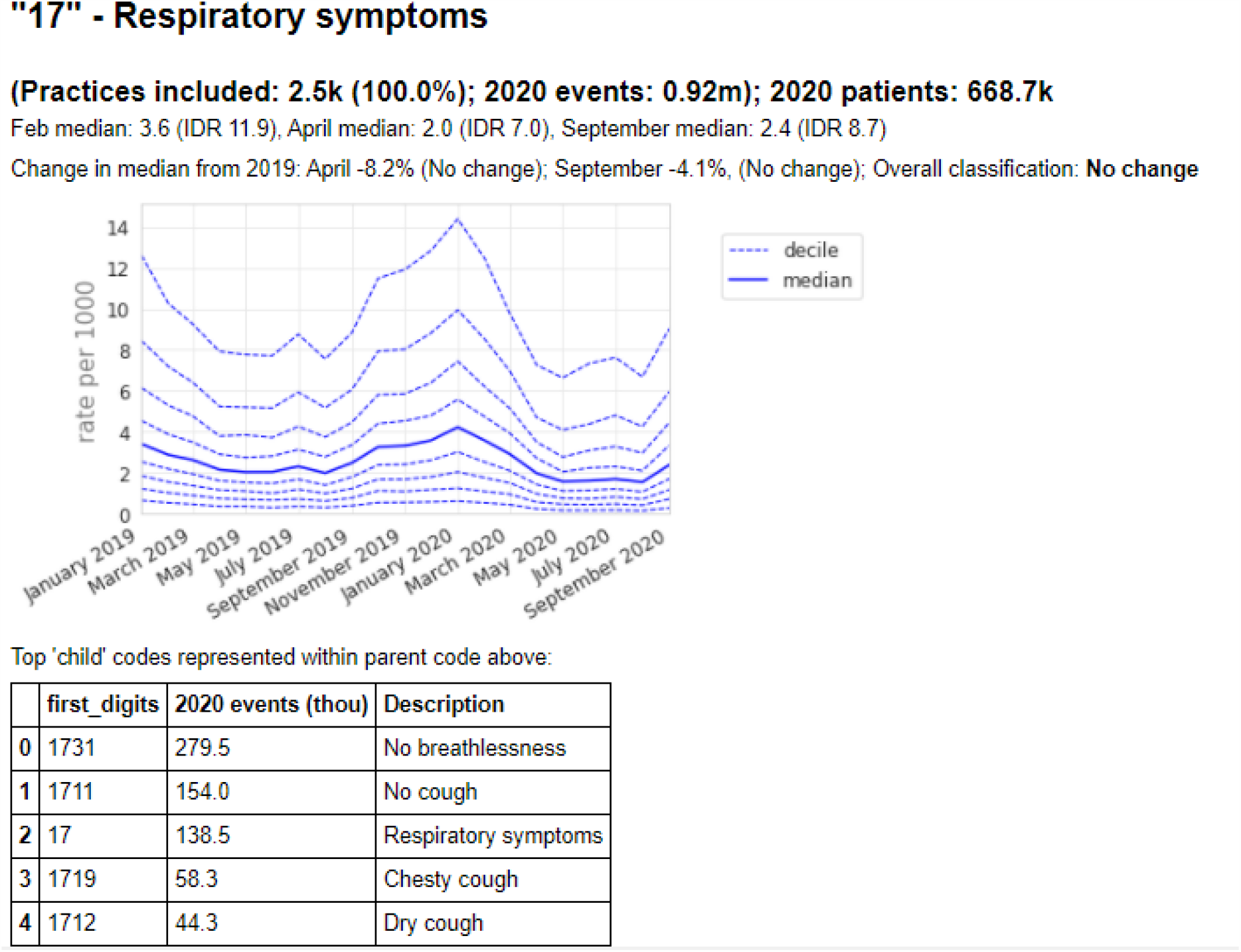
Recording of codes grouped under “Respiratory symptoms” across TPP practices in England (January 2019 - September 2020). The group includes CTV3 codes that begin with “17” and is not necessarily an exhaustive collection of every activity related to Respiratory symptoms. The top 5 codes represented within this group are listed under the graph.

**Figure 5:**
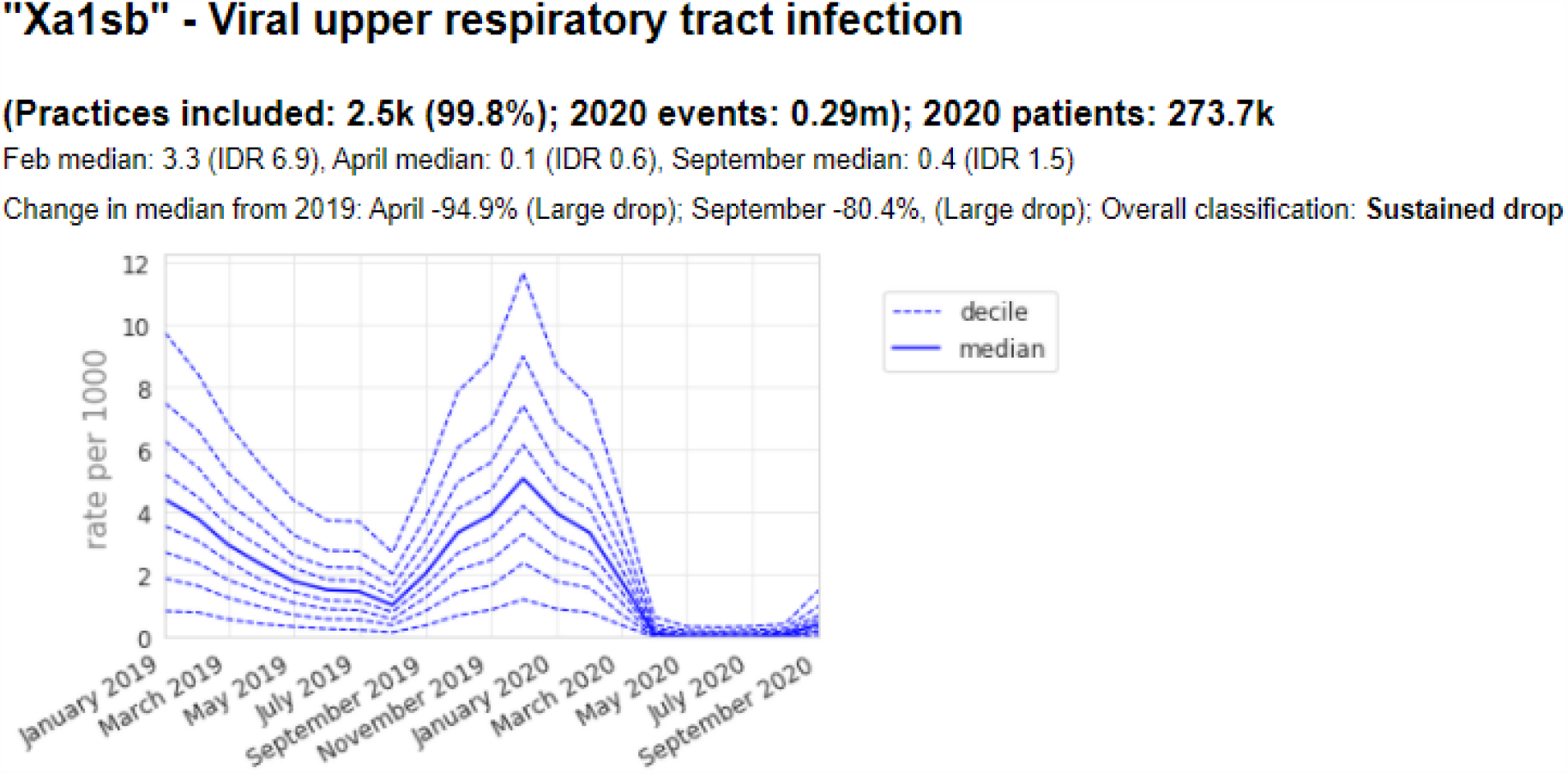
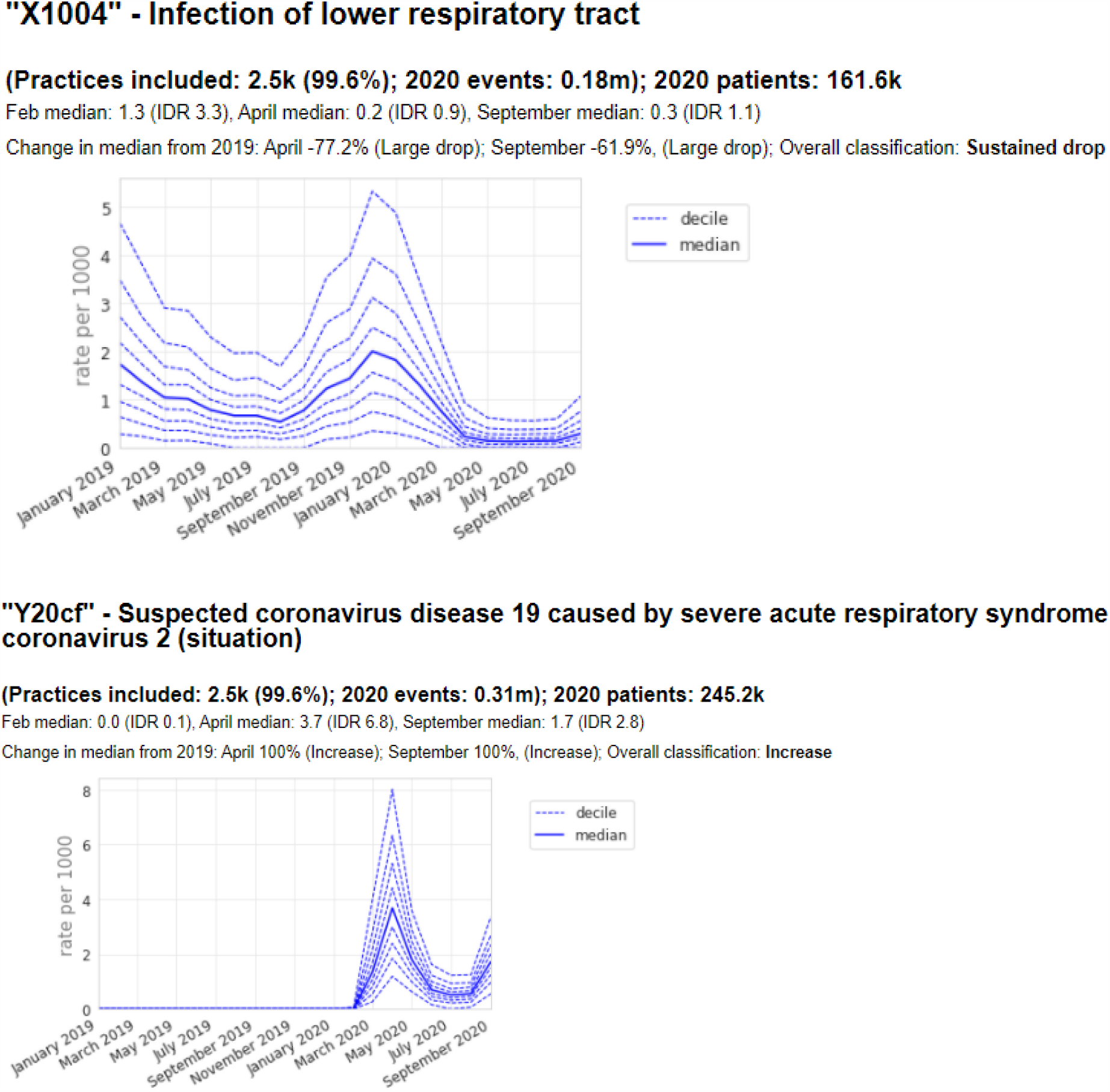
Recording of selected individual codes related to respiratory infections across TPP practices in England (January 2019 - September 2020): (a) Viral upper respiratory tract infection, (b) infection of lower respiratory tract, and (c) Suspected coronavirus disease 19. These were the most common codes we identified for these activities, but other codes may also be used to record the same or similar activities.

### Long-term respiratory conditions

Codes for both “Asthma” and “Chronic obstructive lung disease” were included within the high-level group of codes beginning *“H3”, Chronic obstructive lung disease*, of which 260k codes were recorded in 2020 by 99.8% (n=2.5k) of practices (figure 6). Overall, this group showed a small drop in April (median 0.5/1000, −49.0%) with some recovery by September (median 0.7/1000, −35.7% compared to previous September; figure 6). There were similar trends in some individual codes related to monitoring of these conditions, such as *COPD review* (“Xalet”, figure 7b), with a large drop followed by partial recovery (April −66.7%, September −36.1%). Following a broadly similar pattern, *Asthma annual review* (“Xaleq”, figure 7a) exhibited a smaller drop (April median 2.8/1000, −29.3% from previous April) but with widening variation (IDR 10.6, compared to 7.8 in February with median 5.1/1000). This code had a more complete recovery, to a median of 3.8 in September (−11.8% from the previous September), but with wide variation persisting (IDR 8.4). Several other codes related to asthma monitoring had a gradually increasing pattern, with a dramatic increase in September, including *Asthma control test* (“XaQHq”, figure 7c), *Number of asthma exacerbations in past year* (“XaINh”; Appendix), and *Asthma self-management plan review* (“XaYZB”; Appendix). *Asthma control test*, previously a relatively unusual code but with high variation (February median 0.8/1000, IDR 7.4) rose sharply in September to a median of 2.8/1000 (IDR 10.2, figure 7c).

**Figure 6:**
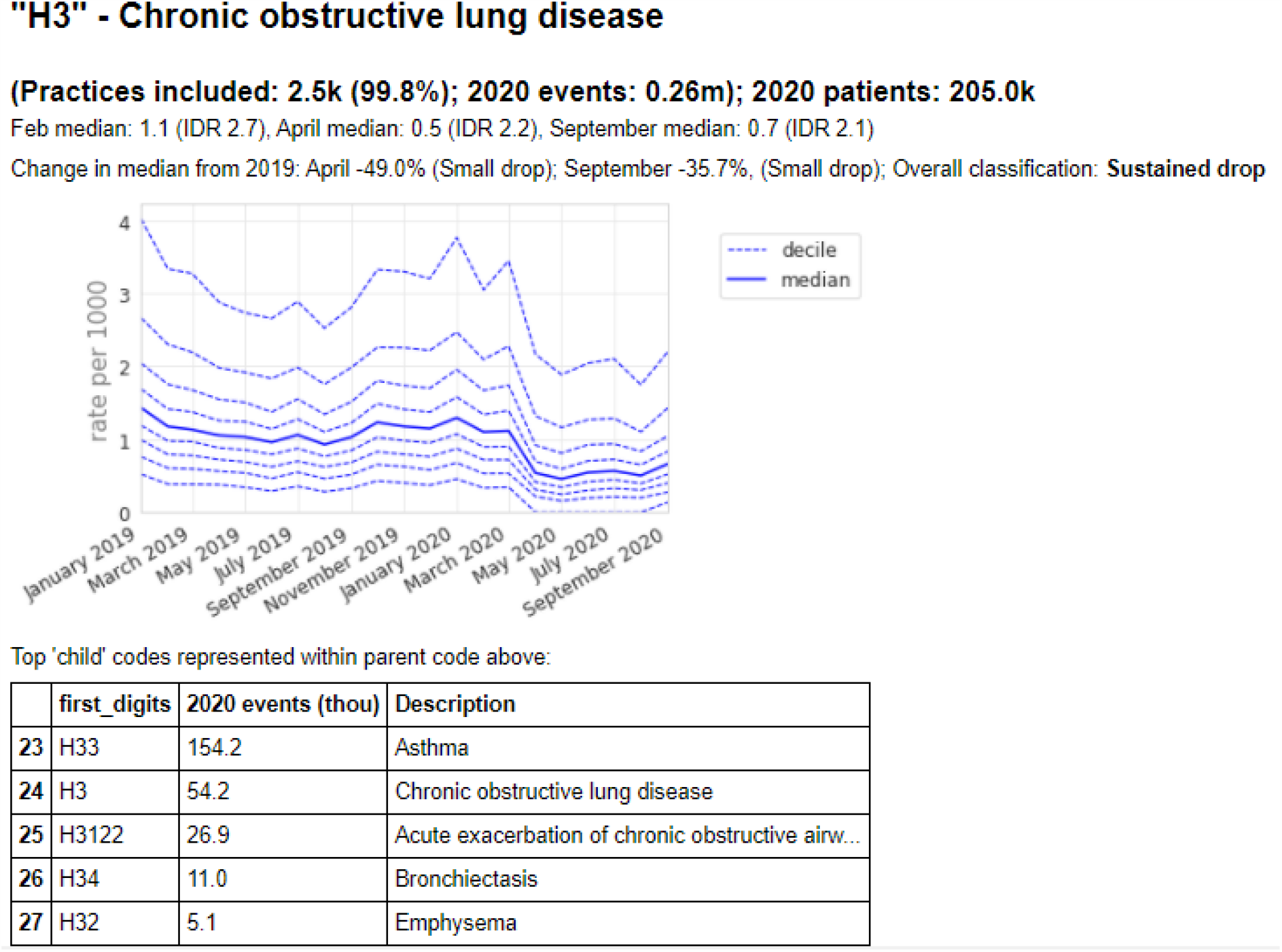
Recording of codes grouped under “Chronic obstructive lung disease” across TPP practices in England (January 2019 - September 2020). The group includes CTV3 codes that begin with “H3” and is not necessarily an exhaustive collection of every activity related to Chronic obstructive lung disease. The top 5 codes represented within this group are listed under the graph.

**Figure 7:**
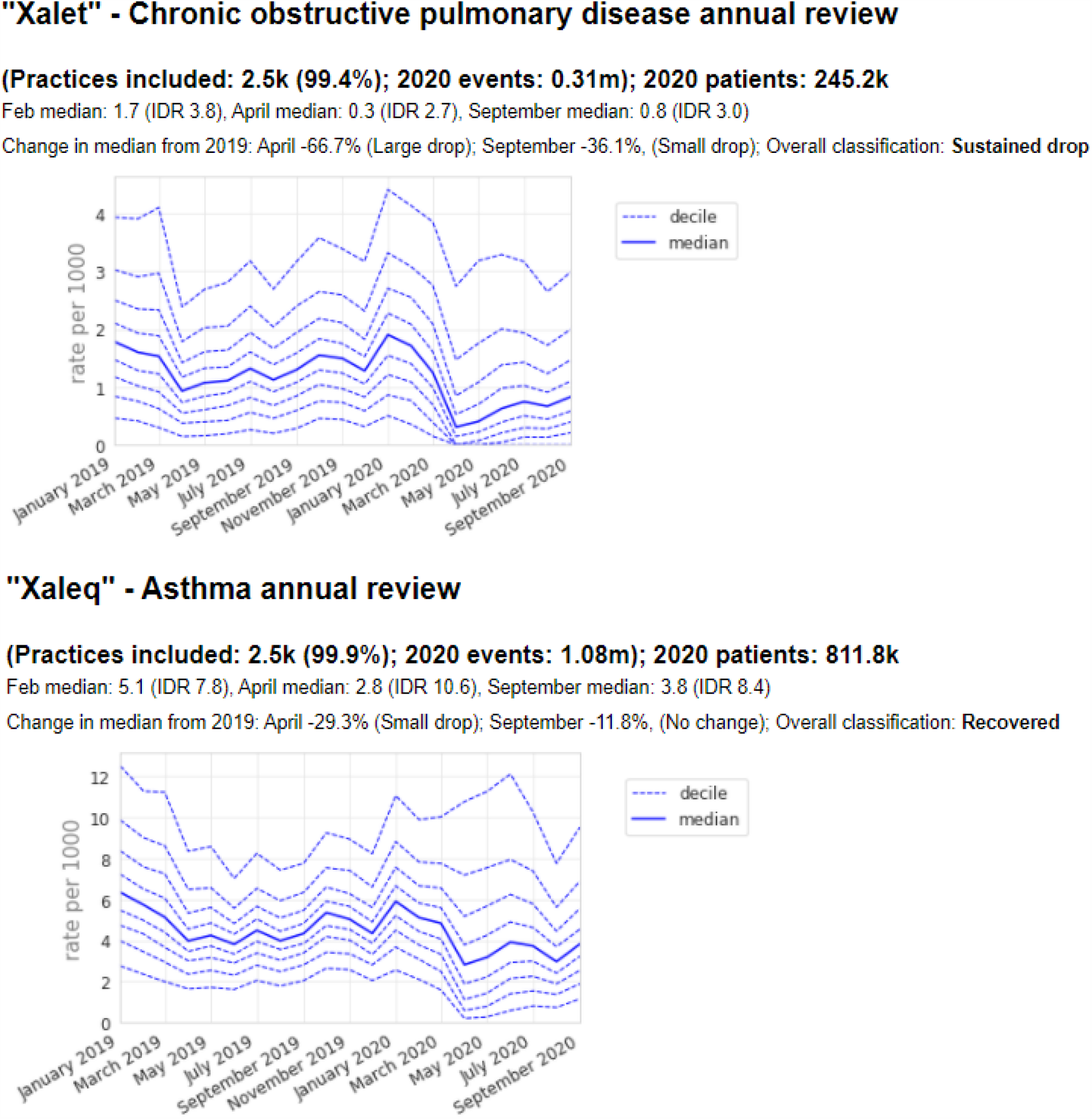
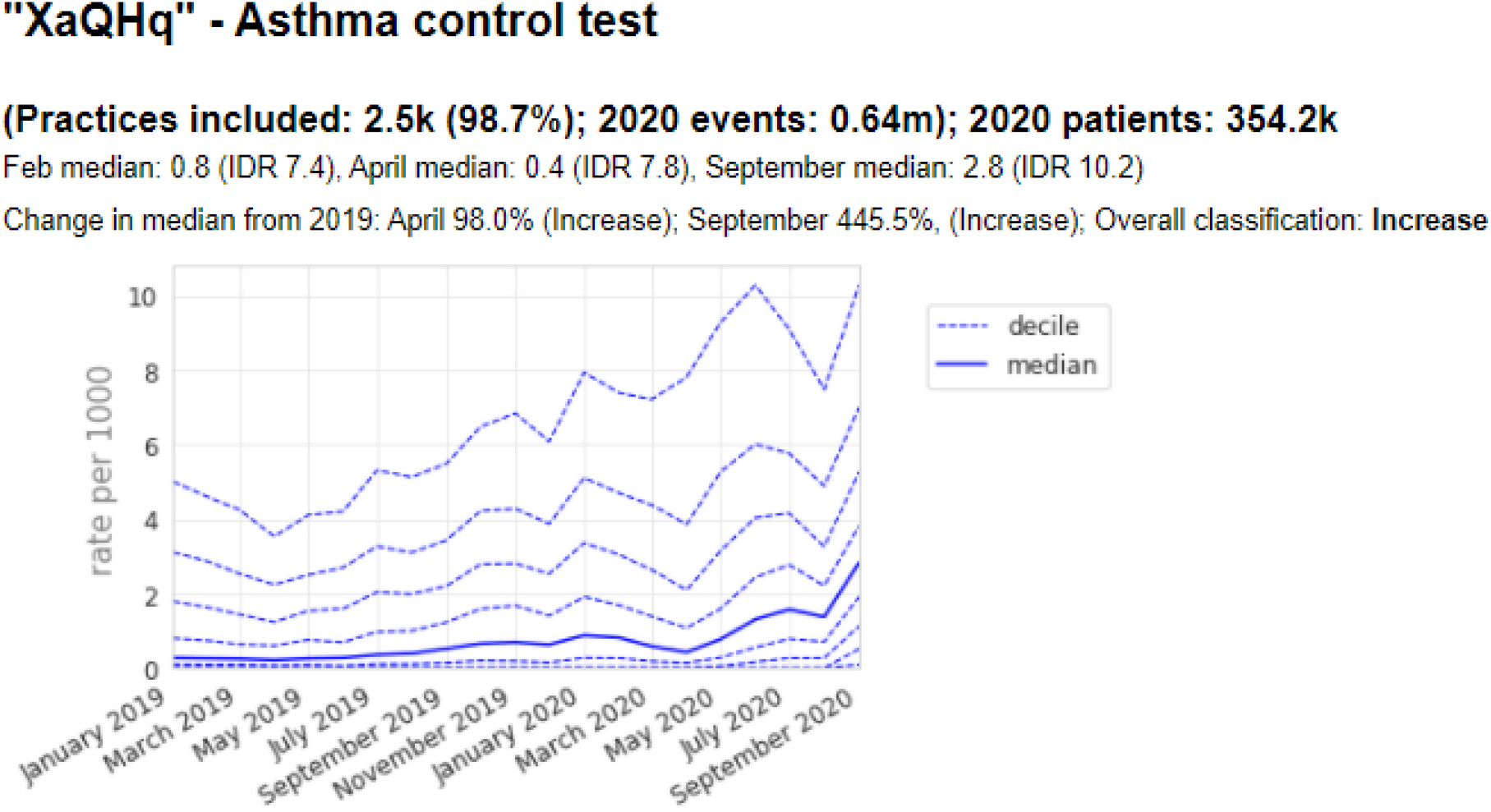
Recording of selected individual codes related to annual reviews for long term respiratory conditions across TPP practices in England (January 2019 - September 2020): (a) COPD annual review, (b) Asthma annual review, (c) asthma control test. These were the most common codes we identified for these activities, but other codes may also be used to record the same or similar activities.

## Discussion

### Summary

We were able to successfully generate data on trends and variation in clinical activity across the records of 40% of all practices in England. We identified substantial and widespread changes in the pattern of clinical activity for laboratory procedures and respiratory disease in primary care since the onset of the COVID-19 pandemic. Broadly, clinical activity related to laboratory procedures such as blood tests declined substantially after “lockdown” but recovered quickly over the summer. However, blood tests to manage high-risk anticoagulants were prioritised and did not experience a similar drop off in activity.

Recording of respiratory symptoms overall, including cough and breathlessness, remained relatively constant, however codes related to viral respiratory illness substantially declined compared to previous activity, with the exception of those specifically related to COVID-19. We observed a small decline associated with high level codes for long-term respiratory conditions such as COPD and asthma. Activity related to asthma annual reviews experienced a small drop but has since recovered, while COPD annual reviews are still below baseline.

### Strengths and weaknesses

The key strengths of this study are the scale and completeness of the underlying raw EHR data. The OpenSAFELY platform runs analyses across an unprecedented scale of data: the full dataset of all raw, single-event-level clinical events for all patients at 40% of all GP practices in England, including all tests, treatments, diagnostic events, and other salient clinical and demographic information for 23.8 million patients. By contrast the CPRD dataset contains records for a substantially smaller number of current patients spread across two databases; and the GPES dataset held by NHS Digital contains a much smaller amount of data on each individual patient. OpenSAFELY also provides data in near-real time, providing unprecedented opportunities for audit and feedback to rapidly identify and resolve concerns around health service activity: the delay from occurrence of a clinical event to it appearing in the OpenSAFELY platform varies from two to nine days. This is substantially faster than any other source of GP data, including those giving much less complete records. It is also faster than any source of large scale secondary care data: the SUS dataset, containing duration and main activity for each hospital event, is coded several weeks after completion (not commencement) of the clinical episode or spell.

We also recognise some limitations. We used a data-driven approach, capitalising on the historical CTV3 hierarchy where possible, in combination with the CTV3 concept hierarchy. There may be other codes relevant to laboratory procedures and respiratory conditions that we have omitted; we intend to iterate our approach with manual curation of “codelists” and utilisation of SNOMED CT hierarchies to better understand activity. With the exception of a small amount of legally restricted data, all occurrences of codes are included, and they do not necessarily indicate unique or new events; for example one patient encounter could generate several similar codes, one patient might have similar diagnoses recorded multiple times over time, or practices might bulk-import information. We studied coding activity and some apparent changes may represent changes in coding behaviour. The large population covered here is likely to be broadly representative of the whole of England’s population, but some coding practices may vary between different EHR systems, so not all of our findings will be generalisable. Finally, codes were counted against patients, who were then allocated to their latest registered practice as at the end of the study period. All patients with an active registration at the end of the study were included, so past activity for patients who registered during the study period was included under their latest practice, even where it occurred in a non-TPP practice. A very small number of patients may have overlapping registrations, meaning any activity they had will be counted against multiple practices. Patients who died or deregistered from TPP practices throughout the study period were not included. Overall, activity counts were up to 6-8% lower than database totals in the earliest months of the study period.

### Findings in Context

A recent systematic review of healthcare usage during the pandemic, encompassing 81 studies across 20 countries, found that healthcare utilisation reduced by approximately one third during the pandemic [11]. This is in line with our findings of substantial reductions in April and May. However, by using near real time data we were also able to detect ongoing recovery. The WHO also found significant disruption to countries’ healthcare capacity for NCDs in May [5] and highlighted the importance that countries *“build back better*” healthcare services for people with NCD, partially as they are more likely to suffer adverse outcomes from COVID-19 [7]. A recent study, not yet peer reviewed, was conducted in the UK Clinical Practice Research Datalink (CPRD) which covers 13% of the UK population: it assessed primary care contacts for specific clinical conditions such as depression, self harm, diabetic emergencies and COPD/asthma exacerbations [12] and found substantial reductions in primary care contacts for acute physical and mental conditions with “limited recovery” by July 2020; similarly we found recovery had occurred in a broad range of coded activity by September. In a separate analysis we found there to be no drop off in INR activity once adjusted for the amount of people on warfarin, the medicine which requires routine INR monitoring [13].

### Future Research

Our study is the first operational research output on its scale using NHS GP data, and to our knowledge also the first to take a primarily data-driven approach to understanding changes in general practice. We propose two broad areas for further research. Firstly building on our approach, a programme of research describing the changes in healthcare activities across a broad range of clinical areas. Secondly, in order to support WHO and NHS England recommendations to “*build back better*”, it is necessary to understand the causes of the changes observed. For example, determining genuine changes in disease prevalence or presentation, versus changes in delivery of healthcare services, or changes in coding behaviour. As an immediate first step we have established a clinical advisory group comprised of: general practitioners, pharmacists, relevant specialists, national clinical advisors with patient and public involvement being coordinated by the Academy of Medical Research Charities. This group will review similar data to those published in this report on a diverse range of subject areas such as; mental health, cardiovascular disease and diabetes. We will synthesise feedback and openly share short written reports describing our findings, identifying any important signals, actionable insights that are suitable for interactive dashboard candidates and research recommendations. We will then actively seek community feedback to iterate our reports. Ultimately, working closely with EHR providers, we aim to present actionable data insights directly back to individual practices to improve patient care and inform response to COVID-19.

### Policy Implications and Interpretation

The COVID-19 pandemic has brought new challenges for the NHS to deliver safe and effective routine care. Our study has shown sharp changes in delivery of activity related to clinical care with quick recovery observed in certain activities. Whilst some important blood tests remained relatively stable throughout the period, most pathology test activity experienced substantial reductions, largely recovering to near-normal levels by September. This may be an indicator of an effective general practice system independently responding in the midst of a global health emergency to deprioritise inessential tests at the height of the pandemic and quickly recovering as the “first wave” subsided. Our proposed *NHS Service Restoration Observatory* can support evaluation of national policies around service restoration and additionally provide opportunities for near real-time audit and feedback to rapidly identify and resolve concerns around health service activity. In particular we hope that data tools such as ours can be used to ensure continuity of high priority clinical services during subsequent waves of the pandemic.

### Summary

We observed substantial changes in activity from April to September 2020 in healthcare service delivery as a result of the COVID-19 pandemic. Whilst some important tests remained relatively stable throughout the period, most pathology test activity experienced substantial reductions, largely recovering to near-normal levels by September. Records of respiratory infections decreased with the exception of codes related to COVID-19, whilst activity of other respiratory disease codes was mixed. The authors are now further developing the *OpenSAFELY NHS Service Restoration Observatory* for real-time monitoring and feedback, using primary care data to measure and rapidly mitigate the indirect health impacts of Covid-19 on the NHS.

## Administrative

## Acknowledgements

We are very grateful for all the support received from the TPP Technical Operations team throughout this work; for close collaboration from all teams running analyses in the OpenSAFELY platform; and for generous assistance and close collaboration from the information governance and database teams at NHS England / NHSX.

## Information governance and ethical approval

NHS England is the data controller; *TPP is the data processor*; and the key researchers on OpenSAFELY are acting on behalf of NHS England. This implementation of OpenSAFELY is hosted within the *TPP environment which is*]accredited to the ISO 27001 information security standard and *is* NHS IG Toolkit compliant;[14,15] patient data has been pseudonymised for analysis and linkage using industry standard cryptographic hashing techniques; all pseudonymised datasets transmitted for linkage onto OpenSAFELY are encrypted; access to the platform is via a virtual private network (VPN) connection, restricted to a small group of researchers; the researchers hold contracts with NHS England and only access the platform to initiate database queries and statistical models; all database activity is logged; only aggregate statistical outputs leave the platform environment following best practice for anonymisation of results such as statistical disclosure control for low cell counts.[16] The OpenSAFELY research platform adheres to the data protection principles of the UK Data Protection Act 2018 and the EU General Data Protection Regulation (GDPR) 2016. In March 2020, the Secretary of State for Health and Social Care used powers under the UK Health Service (Control of Patient Information) Regulations 2002 (COPI) to require organisations to process confidential patient information for the purposes of protecting public health, providing healthcare services to the public and monitoring and managing the COVID-19 outbreak and incidents of exposure; this sets aside the requirement for patient consent.[17] Taken together, these provide the legal bases to link patient datasets on the OpenSAFELY platform. GP practices, from which the primary care data are obtained, are required to share relevant health information to support the public health response to the pandemic, and have been informed of the OpenSAFELY analytics platform.

This study was approved by the Health Research Authority (REC reference 20/LO/0651) and by the LSHTM Ethics Board (reference 21863).

## Guarantor

BG is guarantor.

## Contributorship

*All authors contributed to and approved the final manuscript*.

## CREDIT roles overview

| Contributor Role | Role Definition |
| --- | --- |
| Conceptualization | HC, BMK, BG. |
| Data Curation | HC, JC, CB, SB, PI, DE, SD, GH |
| Funding Acquisition | BG, LS |
| Investigation | HC, PI, CB, BMK, BG, AW, RC, CM, AM |
| Methodology | HC, BG, PI, BMK, CB |
| **Project Administration** | HC, BMK, JM, AM |
| **Resources** | SB, CB, BG, FH |
| **Software** | SB, JC, PI, DE, SD, GH, HC, SH |
| **Supervision** | BG |
| **Visualization** | HC |
| **Writing – Original Draft Preparation** | HC, BMK, BG, JM, AM |
| **Writing – Review & Editing** | All authors |

## Appendix

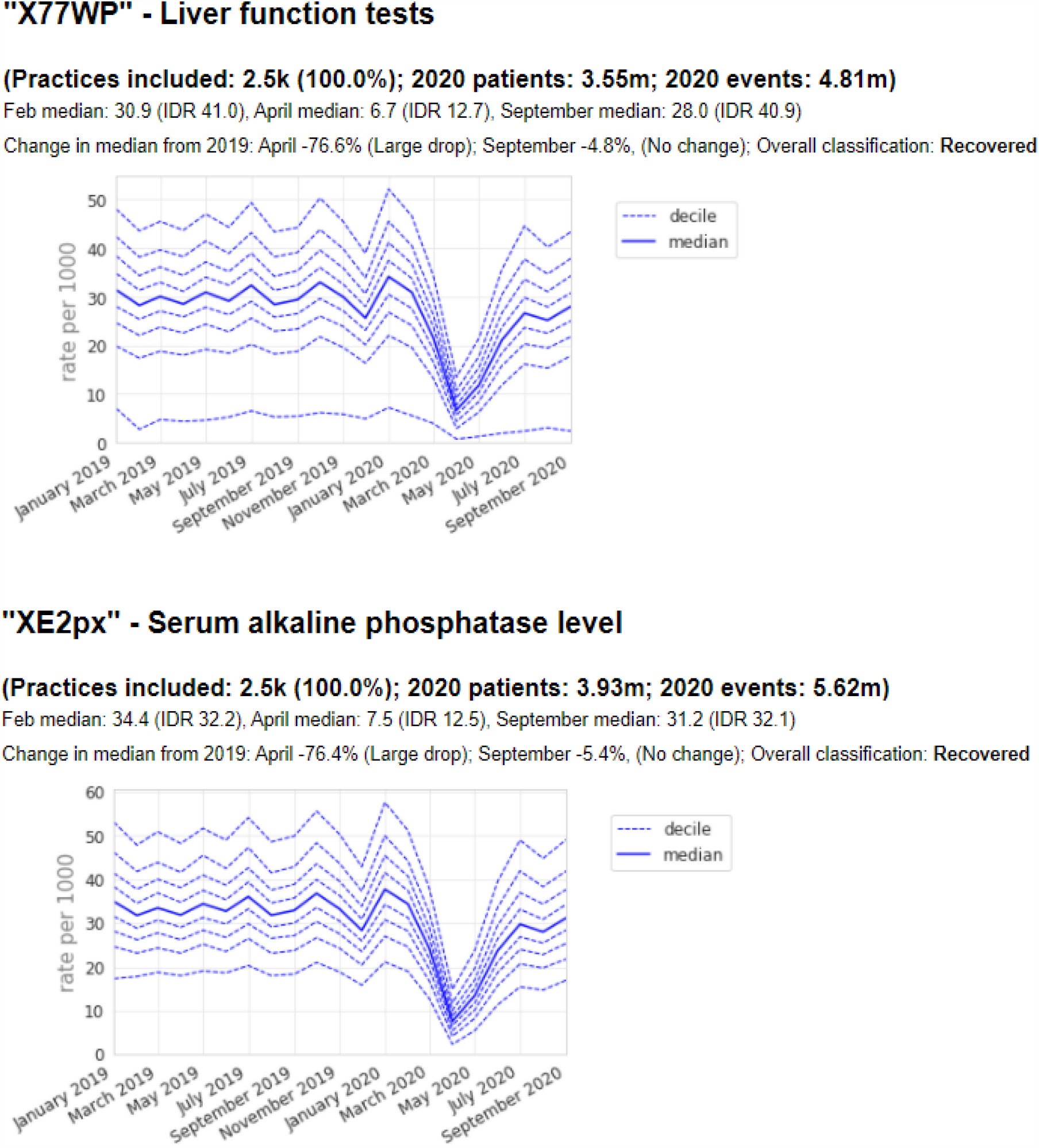
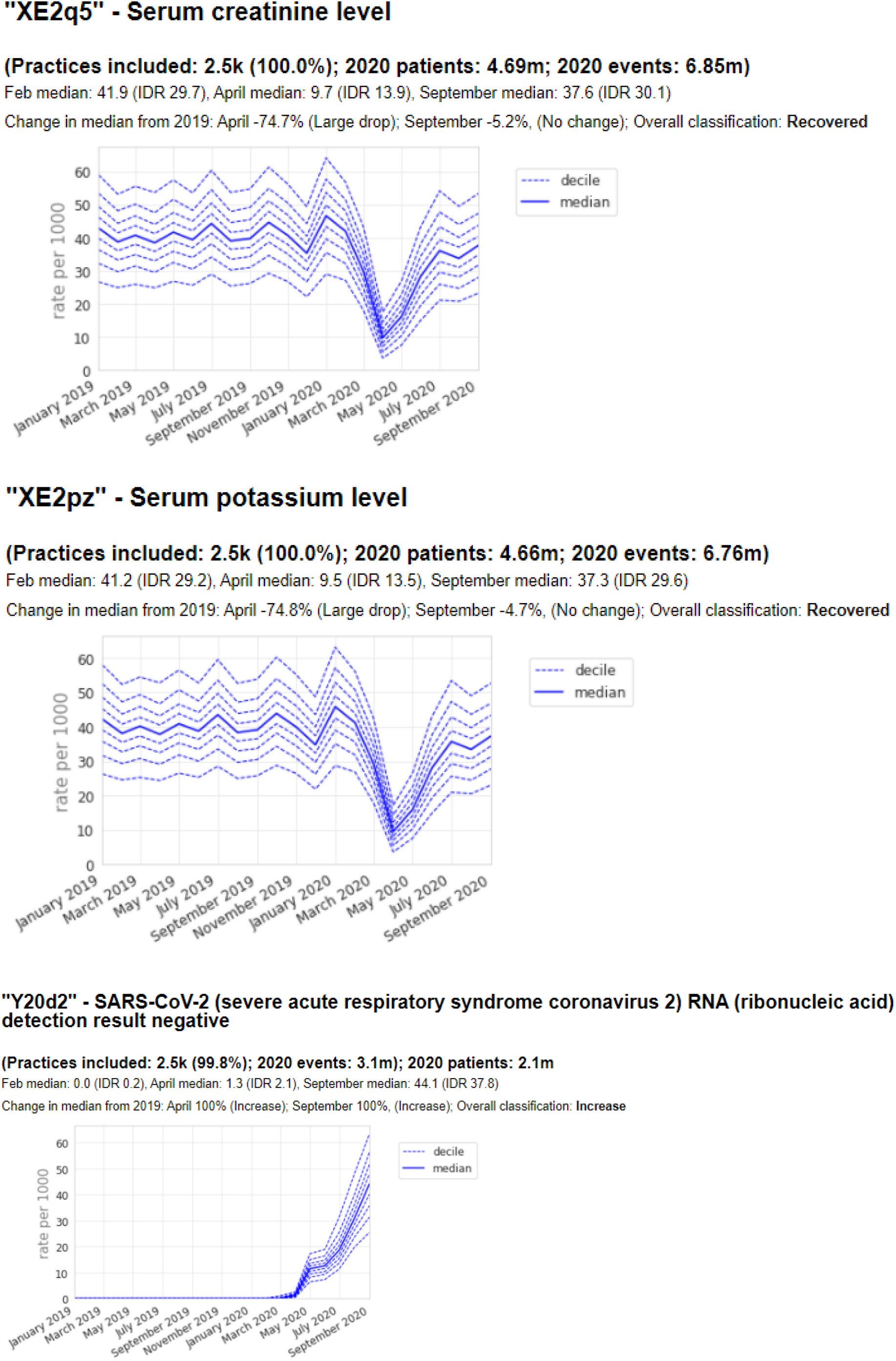
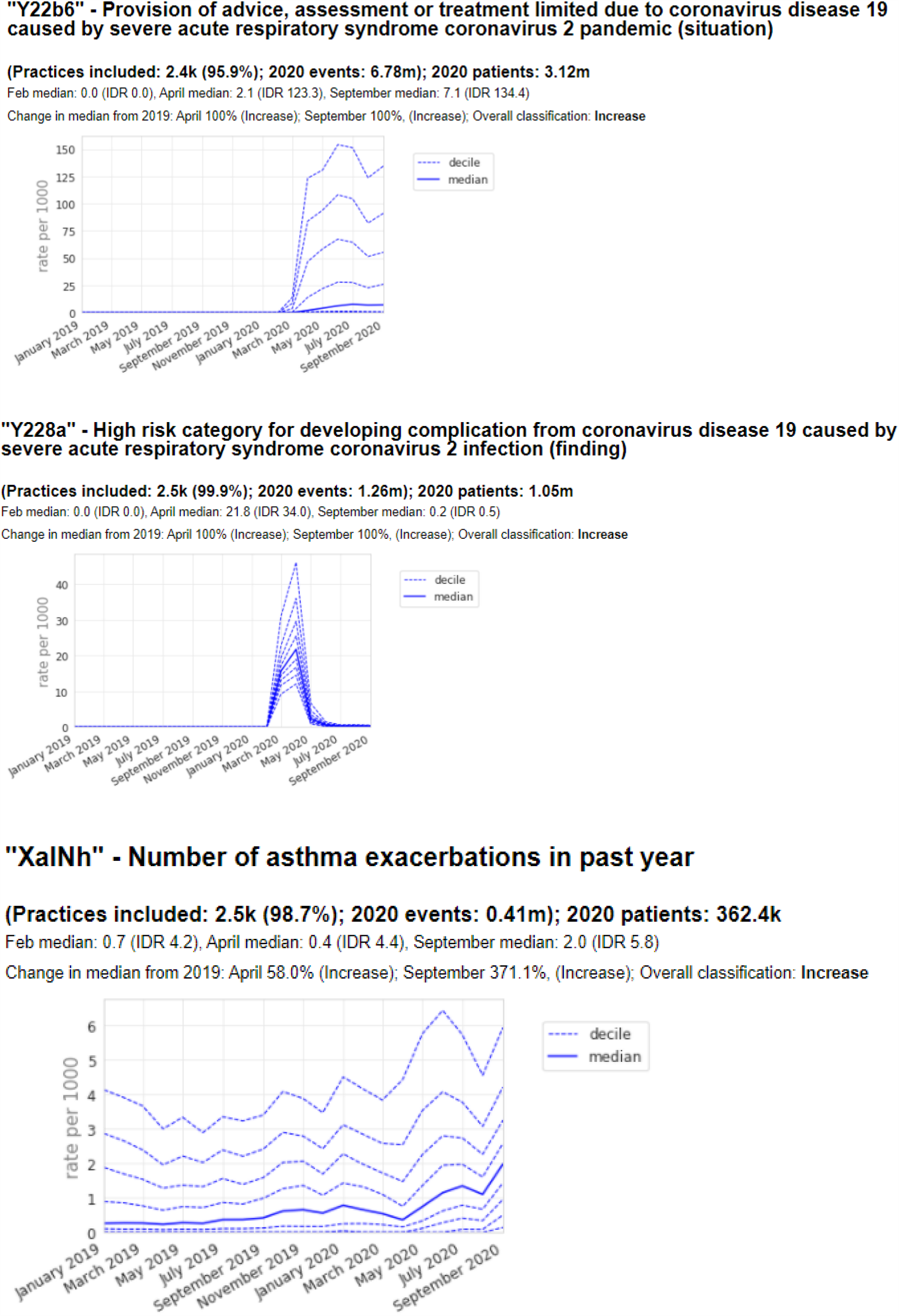
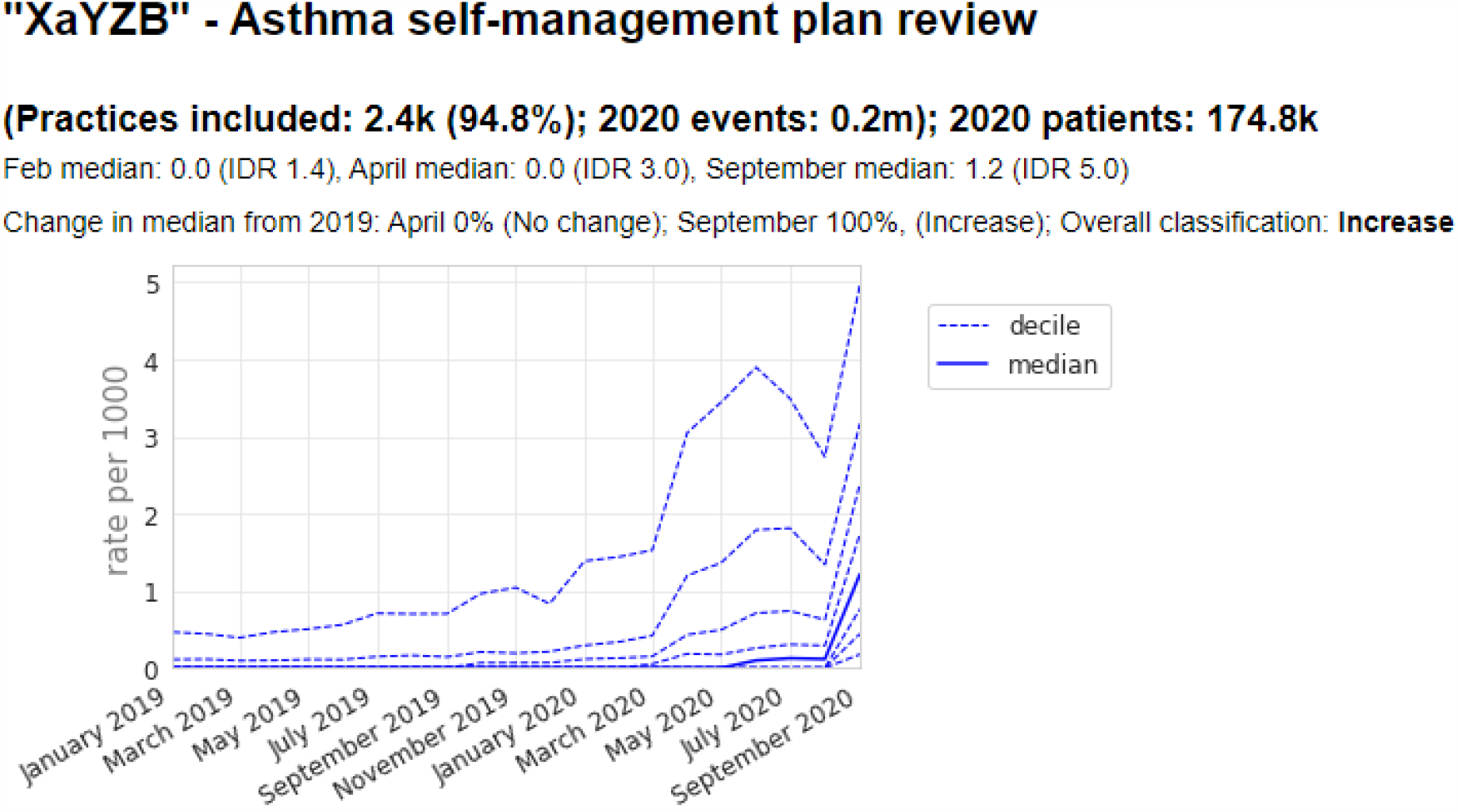

## References

1 COVID-19 situation update worldwide, as of week 50 2020. https://www.ecdc.europa.eu/en/geographical-distribution-2019-ncov-cases(accessed18 Dec 2020).

2 WHO releases guidelines to help countries maintain essential health services during the COVID-19 pandemic. https://www.who.int/news/item/30-03-2020-who-releases-guidelines-to-help-countries-maintain-essential-health-services-during-the-covid-19-pandemic (accessed 12 Nov 2020).

3 WHO. OPERATIONAL PLANNING GUIDELINES TO SUPPORT COUNTRY PREPAREDNESS AND RESPONSE. https://www.who.int/docs/default-source/coronaviruse/covid-19-sprp-unct-guidelines.pdf?sfvrsn=81ff43d8_4

4 NHS England. Urgent next steps on NHS response to covid-19: Simon Stevens letter to local NHS organisations. 2020. https://www.england.nhs.uk/coronavirus/wp-content/uploads/sites/52/2020/03/urgent-next-steps-on-nhs-response-to-covid-19-letter-simon-stevens.pdf

5 Noncommunicable diseases and COVID-19. World Health Organisation. https://www.who.int/teams/noncommunicable-diseases/covid-19 (accessed 11 Nov 2020).

6 NHS England. IMPORTANT – FOR ACTION – THIRD PHASE OF NHS RESPONSE TO COVID-19. 2020. https://www.england.nhs.uk/coronavirus/wp-content/uploads/sites/52/2020/07/20200731-Phase-3-letter-final-1.pdf

7 Williamson EJ, Walker AJ, Bhaskaran K, et al. Factors associated with COVID-19-related death using OpenSAFELY. Nature 2020; 584: 430–6. doi: 10.1038/s41586-020-2521-4

8 OpenSAFELY. https://opensafely.org/ (accessed 25 Nov 2020).

9 NHS Digital. TPP SystmOne: Guide to SNOMED CT changes for General Practice Users. NHS Digital - Kahootz. https://hscic.kahootz.com/gf2.ti/f/762498/43189925/PDF/-/NHS_Digital_SNOMED_CT_in_SystmOne_Short_Guide.pdf: Archive at https://web.archive.org/save/https://hscic.kahootz.com/gf2.ti/f/762498/43189925/PDF/-/NHS_Digital_SNOMED_CT_in_SystmOne_Short_Guide.pdf

10 OpenSAFELY. OpenSAFELY NHS Restoration Observatory Introduction Repo. Github https://github.com/opensafely/restoration-observatory-intro-notebook (accessed 6 Jan 2021).

11 Moynihan R, Sanders S, Michaleff ZA, et al. Pandemic impacts on healthcare utilisation: a systematic review. medRxiv Published Online First: 2020. https://www.medrxiv.org/content/10.1101/2020.10.26.20219352v1.abstract

12 Mansfield KE, Mathur R, Tazare J, et al. COVID-19 collateral: Indirect acute effects of the pandemic on physical and mental health in the UK. Published Online First: 2020. https://papers.ssrn.com/sol3/papers.cfm?abstract_id=3719882

13 Curtis HJ, MacKenna B, Walker AJ, et al. OpenSAFELY: impact of national guidance on switching from warfarin to direct oral anticoagulants (DOACs) in early phase of COVID-19 pandemic in England. medRxiv Published Online First: 2020. https://www.medrxiv.org/content/10.1101/2020.12.03.20243535v1.full

14 BETA – Data Security Standards - NHS Digital. NHS Digital. https://digital.nhs.uk/about-nhs-digital/our-work/nhs-digital-data-and-technology-standards/framework/beta---data-security-standards (accessed 30 Apr 2020).

15 Data Security and Protection Toolkit - NHS Digital. NHS Digital. https://digital.nhs.uk/data-and-information/looking-after-information/data-security-and-information-governance/data-security-and-protection-toolkit (accessed 30 Apr 2020).

16 ISB1523: Anonymisation Standard for Publishing Health and Social Care Data - NHS Digital. NHS Digital. https://digital.nhs.uk/data-and-information/information-standards/information-standards-and-data-collections-including-extractions/publications-and-notifications/standards-and-collections/isb1523-anonymisation-standard-for-publishing-health-and-social-care-data (accessed 30 Apr 2020).

17 Secretary of State for Health and Social Care - UK Government. Coronavirus (COVID- 19): notification to organisations to share information. 2020. https://web.archive.org/web/20200421171727/https://www.gov.uk/government/publications/coronavirus-covid-19-notification-of-data-controllers-to-share-information

